# Method versatility in RNA extraction-free PCR detection of SARS-CoV-2 in saliva samples

**DOI:** 10.1101/2021.12.27.21268334

**Authors:** Orchid M. Allicock, Devyn Yolda-Carr, Rebecca Earnest, Mallery I. Breban, Noel Vega, Isabel M. Ott, Chaney Kalinich, Tara Alpert, Mary E. Petrone, Anne L. Wyllie

**Author notes:** Correspondence: Orchid M. Allicock, 60 College Street, New Haven, CT 06510.

## Abstract

Early in the pandemic, a simple, open-source, RNA extraction-free RT-qPCR protocol for SARS-CoV-2 detection in saliva was developed and made widely available. This simplified approach (SalivaDirect) requires only sample treatment with proteinase K prior to PCR testing. However, feedback from clinical laboratories highlighted a need for a flexible workflow that can be seamlessly integrated into their current health and safety requirements for the receiving and handling of potentially infectious samples. To address these varying needs, we explored additional pre-PCR workflows. We built upon the original SalivaDirect workflow to include an initial incubation step (95°C for 30 minutes, 95°C for 5 minutes or 65°C for 15 minutes) with or without addition of proteinase K. The limit of detection for the workflows tested did not significantly differ from that of the original SalivaDirect workflow. When tested on de-identified saliva samples from confirmed COVID-19 individuals, these workflows also produced comparable virus detection and assay sensitivities, as determined by RT-qPCR analysis. Exclusion of proteinase K did not negatively affect the sensitivity of the assay. The addition of multiple heat pretreatment options to the SalivaDirect protocol increases the accessibility of this cost-effective SARS-CoV-2 test as it gives diagnostic laboratories the flexibility to implement the workflow which best suits their safety protocols.

## Introduction

Almost two years after the emergence of SARS-CoV-2, diagnostic testing remains an important mitigation strategy. As outbreaks and testing policies evolve and as screening testing has emerged as a key feature enabling communities to safely re-open, labs have had to adapt to the changing needs of their local communities. Throughout, this has often required the rapid implementation of alternative or even novel strategies to meet testing demands. However efforts have been hampered with staffing shortages, supply chain disruptions and slow regulatory approval for alternative test protocols or testing instrumentation. Combined, these challenges highlight a great need for alternative testing strategies that a) utilize locally available and inexpensive testing materials, and b) are easy to adopt in either existing or newly created COVID-19 testing laboratories.

Alternative testing strategies should fit seamlessly into an existing workflow of a laboratory, while adhering to relevant biosafety and biosecurity requirements. Within the limits imposed by the mandatory Laboratory Biosafety Guidelines for Handling and Processing Specimens issued by governing bodies^1^, each laboratory has their own protocols when it comes to the intake and processing of infectious agents. As such, adopting additional protocols to help meet mass testing needs, with limited or no flexibility in the reagents, kits or instrumentation permitted for use, can result in delayed test implementation due to the additional investment required or supply chain disruptions.

In an effort to increase access to COVID-19 testing by minimizing test implementation challenges, we developed a freely available, open-source saliva-based RT-qPCR diagnostic assay (SalivaDirect) with a simplified and flexible workflow. Key to this approach was obviating the need for sample collection by trained healthcare personnel, removing the requirement for specific collection devices and transport media, while validating reagents and instrumentation from multiple suppliers to enable laboratories to utilize their existing infrastructure, or when needed, to help circumvent supply chain disruptions.

However, as SalivaDirect was made available to laboratories around the US, the diversity in specimen handling processing requirements when working with potentially infectious samples containing this novel coronavirus limited implementation in some sites. Upon receipt of clinical samples, laboratories employ different strategies for viral inactivation before processing including the addition of solvent/detergents, low pH inactivation, irradiation, or heat ^2, 3^. Previous studies have demonstrated that heat alone is capable of effective viral inactivation of SARS ^4^ and Middle East Respiratory Syndrome ^5, 6^, and more recently, also for SARS-CoV-2 ^2, 7, 8^. Given the affordability and broad availability of heating sources in clinical laboratories (e.g. heating block/water bath), we sought to explore additional workflows which incorporate heat-pretreatment to permit safer sample handling. Using spiked and clinical saliva samples, we evaluated the effect of thermal incubation of samples on the sensitivity of SARS-CoV-2 detection prior to testing in the SalivaDirect assay.

## Materials and Methods

### Ethics statement and sample collection

For the spiking experiments, the use of de-identified specimens from healthy or SARS-CoV-2-positive individuals was approved by the Institutional Review Board of the Yale Human Research Protection Program (FWA00002571, Protocol ID. 2000027690) ^9^. For the clinical evaluation of the workflows, de-identified saliva samples were collected using previously developed saliva self-collection protocols ^10^, and was approved by the Institutional Review Board of the Yale Human Research Protection Program (Protocol ID. 2000029876). For both studies, participants were informed in writing about the purpose and procedure of the study, and consented to study participation through the act of providing the saliva sample. All samples were transferred at room temperature to the laboratory, and stored within 12 hours at -80°C until further analysis.

### Alternate workflows

Saliva samples were thawed on ice and processed using the seven workflows detailed in Figure 1. Each of the six new workflows (**Figures 1B and 1C**) was compared with the original SalivaDirect protocol (**Figure 1A**). Each sample was first incubated at each of the three different heat pretreatment conditions (65°C for 15 minutes, 95°C for 5 minutes and 95°C for 30 minutes) using a heating block or a thermocycler. After incubation the samples were split into 2 aliquots; 10 μL was stored at 4°C until testing in RT-qPCR, and 50 μL was placed in a separate tube with 2.5 μL (50mg/mL) proteinase K (Thermo Fisher). After vortexing for 1 minute, the samples were incubated at 95°C for 5 minutes to inactivate the proteinase K. Following sample processing, 5 µL of the saliva lysates (either stored at 4°C or treated with proteinase K) were tested using the SalivaDirect real-time RT-qPCR assay ^11^. This assay uses primers and probes from the US CDC, targeting the nucleocapsid gene (N1 2019-nCoV_N1) and the human RNase P (RP) as an extraction control ^12^. The RT-qPCR was performed using the Luna Universal Probe One-Step RT-qPCR Kit (New England Biolabs) on the BioRad CFX96 Touch (BioRad, CA). A synthetic SARS-CoV-2 RNA control from Twist Bioscience (San Francisco, CA) was diluted to 100 copies/µL and used as the positive control for N1.

**Figure 1.**
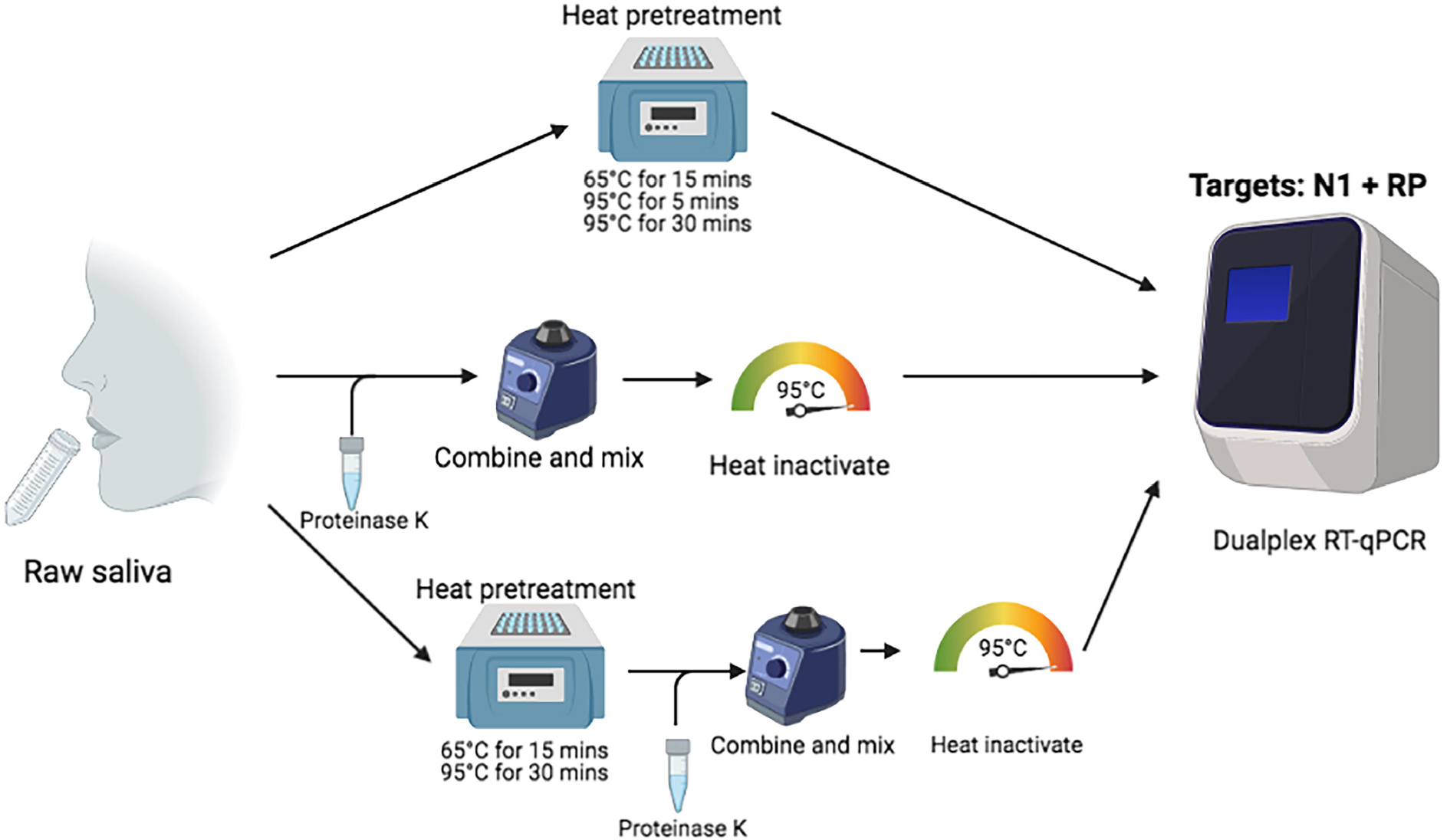
Alternative SalivaDirect workflows including heat-pretreatment prior to sample processing. In the original SalivaDirect protocol, (A) raw saliva is combined with proteinase K then followed by heat-inactivation of the proteinase K and testing in RT-qPCR. The alternate workflows include an initial heat treatment step (65°C for 15 minutes, 95°C for 5 minutes or 95°C for 30 minutes), followed by either (B) testing by RT-qPCR directly or (C) the addition and inactivation of proteinase K prior to testing by RT-qPCR. (Figure created with BioRender.com)

### Limit of detection

We performed a limit of detection confirmation study to evaluate the sensitivity of SARS-CoV-2 detection when testing samples with a heat-pretreatment step. Samples were prepared by spiking SARS-CoV-2-positive saliva from a healthcare worker diagnosed with COVID-19 with a known virus concentration (3.7 × 10^4^ copies/µL) into saliva samples from healthy individuals (negative for SARS-CoV-2 RNA). Spiked saliva samples at concentrations of 50, 25 and 12 copies/µL were tested in triplicate, and concentrations of 6, 3, and 1.5 copies/µL were tested with 20 individual replicates. All samples were tested using the seven workflows depicted in **Figure 1**. The limit of detection for each workflow was determined to be the lowest concentration at which at least 19/20 replicates were positive for SARS-CoV-2 (cycle threshold (Ct) value <40.0).

### Workflow validation with SARS-CoV-2 clinical specimens

We validated each of the different workflows using 20 de-identified clinical saliva specimens which previously tested positive for SARS-CoV-2 using the standard SalivaDirect workflow (Figure 1A). Each sample was processed by each of the six workflows, and resulting Ct values were compared to those obtained when originally tested by the standard SalivaDirect protocol.

### Data analysis

The sensitivity of the different workflows were compared using repeated-measures ANOVA on the Ct values of each replicate or isolate for each workflow, followed by a Tukey post-hoc test to compare individual pairs of conditions. The agreement of Ct values between each of the alternate workflows and original SalivaDirect protocol were assessed using Pearson correlation coefficient. To compare the Ct value in the clinical samples a Wilcoxon signed-rank test was performed. The negative RT-qPCR of the target gene was set at the Ct value of 40 for the statistical analysis. A *p* value < 0.05 was considered statistically significant. All analyses were conducted with GraphPad Prism 9.1.2 (GraphPad Software, Inc., San Diego, CA).

## Results

### Limit of detection

The limit of detection (LOD) for the original SalivaDirect protocol (reagents and PCR instrument, depending) is as few as 1.5 copies/μL ^13^, with the formal LOD recorded at 12 copies/μL, reflecting the least sensitive combination of recommended reagents and instruments ^11^. To determine the LOD of the alternate workflows, we tested spiked saliva samples at 50, 25 and 12 virus RNA copies/μL in triplicate, all of which were detected by RT-qPCR (**Table S1**). We confirmed the LOD for each workflow by testing 20 replicates of spiked saliva samples at lower viral copies (6, 3 and 1.5 virus RNA copies/μL). While all 20 of the individual replicates of the spiked samples at 6 virus RNA copies/μL tested positive for each workflow (**Figure 2, Table 1**), some workflows were more sensitive, with workflows including a heat incubation step of 65°C for 15 minutes without proteinase K, and 95°C for 5 minutes with or without proteinase K having a limit of detection of 3 virus RNA copies/μL.

**Figure 2.**
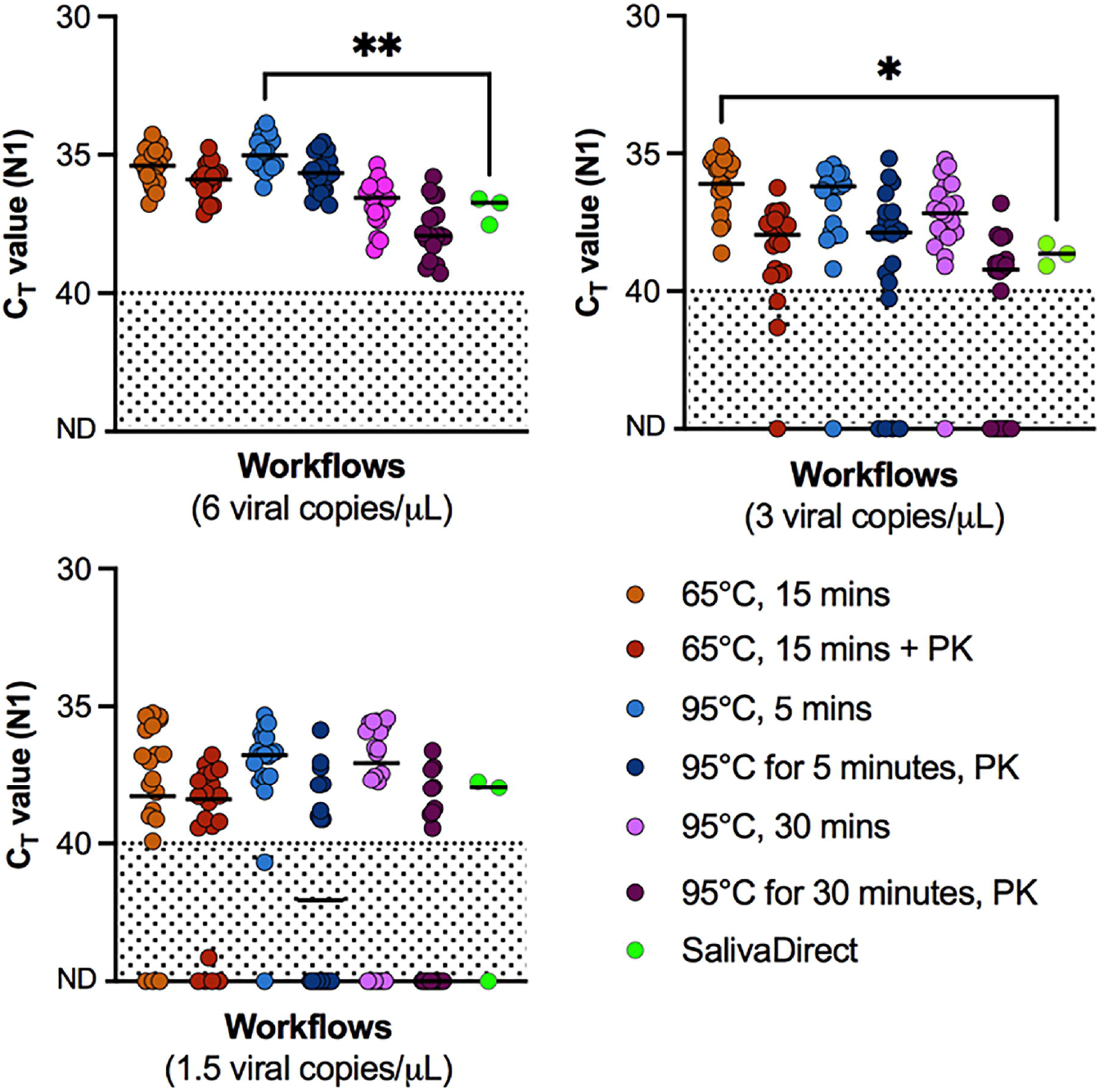
Comparison of the sensitivity of the 6 alternate workflows of the SalivaDirect RT-qPCR saliva-based assay for SARS-CoV-2 detection. Each alternative workflow was evaluated by processing spiked positive saliva at (A) 6, (B) 3, and (C) 1.5 copies/µL in 20 replicates, and comparing the CT values as a proxy for workflow sensitivity. Solid lines indicate the median Ct values targeting the N1 gene. The dashed line indicates the detection limit at 40 Ct with samples falling below the dashed line considered negative for SARS-CoV-2.The differences between each of the workflows and the original SalivaDirect protocol were compared by a Wilcoxon test test (p < 0.05, * = 0.03, ** = 0.001). Data used to make this figure can be found in **Table S1**. *Abbreviations: Ct, cycle threshold; ND, not detected; PK, proteinase K*.

**Table 1.**
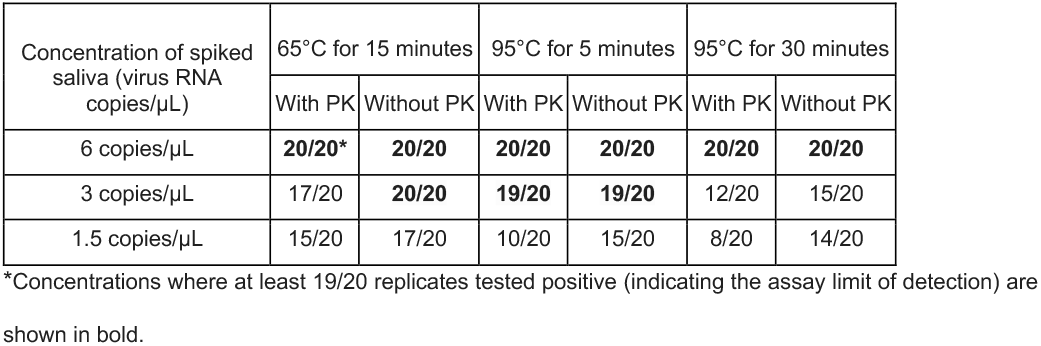
The number of replicates of spiked saliva samples which were considered positive following testing in the alternative workflows including heat pre-treatment with or without treatment with proteinase K (PK).

### Clinical evaluation of heat treatment workflows

To investigate the agreement between each of the different workflows and the original SalivaDirect protocol, 20 saliva samples which previously tested positive for SARS-CoV-2 by SalivaDirect (Ct value range 22.16 -38.71) were processed using each of the different workflows. There was a median Ct difference in workflows of 0.92 to 6.08 across each of the clinical samples (**Figure 3A**). Ct values from the N1 gene obtained from the original saliva direct workflow correlated with each of the alternate workflows (Pearson r > 0.9, *p* <0.0001) (**Figure S1, Table S2**). The workflow that was significantly less efficient at detecting N1 gene than the original was 95°C for 30 minutes with the added proteinase K step (ΔCt = 1.46, *p* < 0.001).

**Figure 3.**
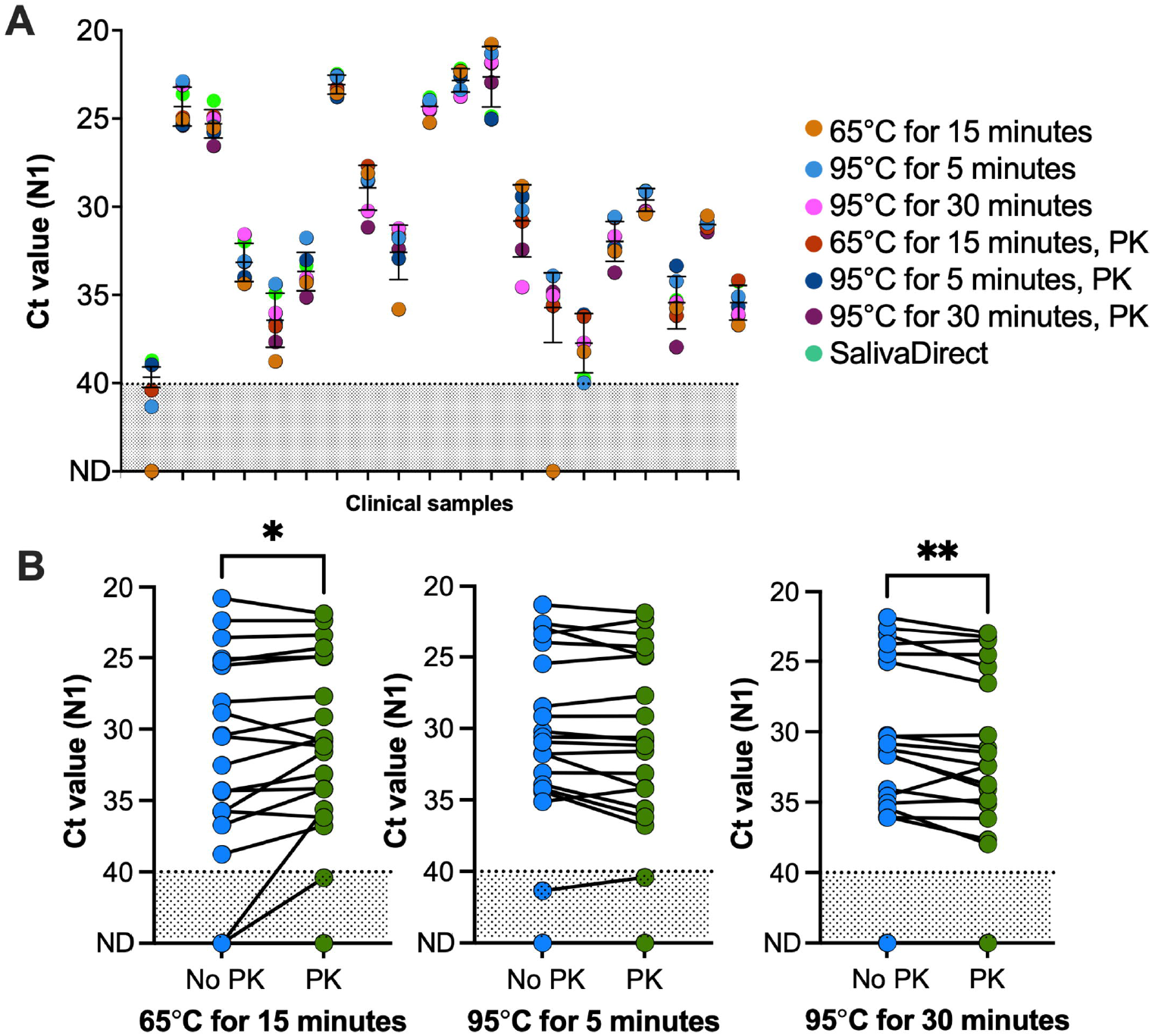
Detection of SARS-CoV-2 RNA in positive clinical saliva samples across the 7 different workflows. Clinical saliva samples positive for SARS-CoV-2 were used to compare the sensitivities in detection across the 7 workflows. (A) Ct values for each of the 20 clinical samples processed with the different workflows. The solid lines indicate the median Ct values and 95% CI for each sample across the different workflows. Each dot represents an individual replicate of each of the 20 clinical samples as indicated by the dash on the x-axis. (B) We evaluated the effect of proteinase K on the sensitivity of detection between each of 3 different heat treatment conditions by a Wilcoxon test (*p* < 0.05). Matched clinical samples are represented by the solid black line between those processed without proteinase K (blue dots), and those with proteinase K (green dots). The area below the dotted line indicates the detection threshold for the N1 gene. Data used to make this figure can be found in Table S2. (*, *p* = 0.027; **, *p* = 0.009) *Abbreviations: Ct, cycle threshold; ND, not detected; PK, proteinase K*.

We also investigated the impact of proteinase K on the sensitivity of each workflow (**Figure 3B**). While the addition of proteinase K made no difference when the samples were first incubated at 95°C for 5 minutes, the addition of proteinase K following incubation at 65°C for 15 minutes resulted in an increase in sensitivity (median difference in Ct value = -0.53, *p* = 0.027). When the proteinase K was added following incubation at 95°C for 30 minutes however, there was a decrease in sensitivity (median difference in Ct value = 0.79, *p* = 0.009).

The use of the human *RNase P* (RP) control gene in the dualplex PCR helps to monitor for any major degradation or inhibitors in samples. Ct values for RP were significantly higher (indicating reduced detection) for all workflows except when the samples were processed with incubation steps of 65°C for 15 minutes with PK, and 95°C for 5 minutes with PK. None of the saliva samples had Ct values for RP over 35, the threshold for an invalid sample (**Table S2**), demonstrating that none of the workflows negatively affected the quality of the clinical samples.

## Discussion

Saliva as a specimen type is underutilized in molecular diagnostics. Prior to 2020, almost all molecular-based diagnostic tests for respiratory infections required nasopharyngeal specimens (e.g aspirate or swabs) with only one test using saliva swabs for the detection of cytomegalovirus ^14^. So following the first reports outlining the potential of saliva as a reliable sample type for SARS-CoV-2 detection ^9, 15, 16^ and a means to overcome the numerous challenges that labs were facing with nasopharyngeal swabs, diagnostic and research laboratories alike scrambled to devise guidelines and protocols for the safe handling, processing and storage of potentially infectious saliva specimens. Heating samples on arrival for virus inactivation presents a straightforward approach which can be easily adapted in a range of laboratory settings. As an additional benefit, heating of samples decreases the viscosity of saliva samples ^17^, making sample pipetting easier.

In the current study we investigated the addition of thermal incubation prior to testing saliva samples with the simplified RT-qPCR assay, SalivaDirect, and evaluated the effect of heat pre-treatment on the sensitivity of SARS-CoV-2 detection. Although 56°C is commonly used for inactivation of enveloped viruses ^18^, higher temperatures have been used for other coronaviruses ^6^. More recently, it has been shown that SARS-CoV-2 viral particles can be inactivated by incubating a sample at 65°C for 15 minutes and 95°C for 5 minutes before processing^7, 8, 19^, without loss of sensitivity in nasopharyngeal swabs and sera ^2, 7^. While in the current study, we did not confirm the inactivation of SARS-CoV-2 following heat pretreatment, the conditions selected were based on the literature demonstrating the total inactivation of SARS-CoV-2, or, in the case of 95°C for 30 minutes (workflow C), at the request of testing laboratories who were already utilizing this step prior to testing by alternative methods.

For all workflows investigated, we found a limit of detection of 3 to 6 copies/μL. The robustness of the detection of the N1 gene in saliva has been demonstrated previously, with no significant decrease in sensitivity when exposed to moderate heat ^10, 20^. Additionally, proteinase K also did not significantly affect the overall sensitivity of detection of the N1 gene (paired t test, *p* = 0.247). Rather, at the lower incubation temperature (65°C for 15 minutes), addition of proteinase K marginally increased the sensitivity of detection as compared to samples processed without proteinase K. It is possible that, while 65°C alone can effectively inactivate SARS CoV-2, this is not as effective at “extracting” all of the virus RNA for RT-qPCR detection. Conversely, the addition of the proteinase K step to samples incubated at 95°C for 30 minutes significantly decreased the sensitivity of both N1 and RNAse P detection when compared to the original SalivaDirect protocol. However, while RP is used as an internal control, the minor differences observed would not affect the outcome of the assay. Proteinase K degrades RNases and assists in preserving RNA integrity, hence it is commonly used in the processing of samples for molecular assays, especially in various extraction-free workflows^21, 22^. Extraction-free workflows with saliva is no exception, and has been incorporated into many saliva-based extraction-free molecular assays ^23-26^, including SalivaDirect.

The addition of alternate workflows to an already flexible testing framework further support the rapid implementation of the SalivaDirect RT-qPCR assay in diagnostic and research laboratories looking to improve access to SARS-CoV-2 testing in their local communities. Importantly, with the addition of pre-treatment heat steps, these expanded workflows help to prevent the exposure of laboratory personnel directly or indirectly handling potentially SARS-CoV-2-infected samples, while providing flexibility of adaptation into their existing standard operating procedures. One of the unique attributes of SalivaDirect is it is validated with materials from multiple vendors, to minimize the risk of supply chain issues. The option of omitting proteinase K treatment makes this flexible framework even less vulnerable to supply shortages, and more affordable for laboratories - and thus importantly, to their patients. This is especially important for the implementation of mass testing strategies in resource-poor areas, or in low-to-middle income countries which continue to suffer the brunt of the reagent and laboratory consumables shortages during the pandemic.

## Data Availability

All data produced in the present study are available upon reasonable request to the authors.

## Data Availability

All of the data generated from this study is in the Supplementary information.

## Author contributions

A.L.W. conceived the study. O.M.A. and R.E. assisted with the coordination and execution of the study. R.E., M.E.P., C.K. and T.A. coordinated sample collection. M.I.B., I.M.O, D.Y-C., O.M.A. and N.V. performed the diagnostic tests. O.M.A. and N.V. analyzed the data. O.M.A. assisted with the design of the statistical analysis. O.M.A., N.V., and A.L.W. wrote and edited the manuscript.

## Acknowledgements

We thank the study participants for their time and cooperation. This work was funded by Tempus Labs, Inc (A.L.W), Yale Center for Clinical Investigation TL1 TR001864 (M.E.P.) and Fast Grant from Emergent Ventures at the Mercatus Center at George Mason University (A.L.W).

