## Supplementary figures and images for "Method versatility in RNA extraction-free PCR detection of SARS-CoV-2 in saliva samples"

### Supplemental figure 1

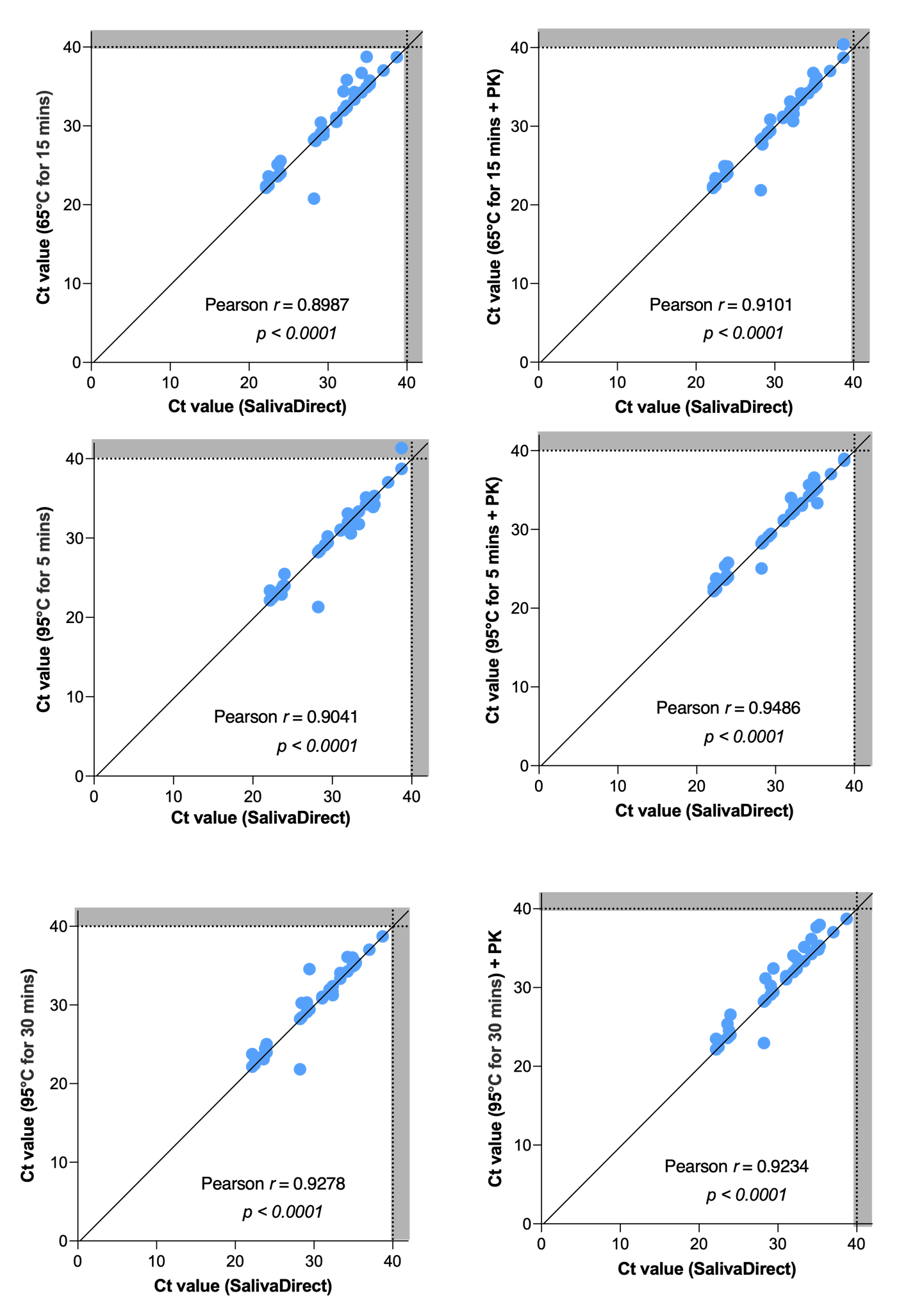
